# Social Determinants of In-Hospital AIDS-Related Mortality: A Cohort Study of 13,266 Low-Income Patients in Brazil

**DOI:** 10.64898/2026.02.26.26347160

**Authors:** Priscila Scaff Gestal, Gabriela S Jesus, Andrea F Silva, Iracema Lua, Carlos AS Teles, Luis E Souza, James Macinko, Inês Dourado, Davide Rasella

## Abstract

**Background:** Social determinants of health (SDOH)—including socioeconomic, demographic, and geographic factors—are closely linked to HIV/AIDS outcomes. This study assessed the association between SDOH and in-hospital AIDS-related mortality among low-income individuals in Brazil.

**Methods:** We conducted a cohort study using data from the 100 Million Brazilian Cohort, which includes individuals who applied for government social programs (2008–2018), linked to national HIV/AIDS hospitalization and mortality records. Multivariable Poisson regression with a hierarchical framework was used to evaluate associations between individual and household-level SDOH and in-hospital AIDS mortality.

**Findings:** Among 13,266 individuals hospitalized for AIDS, 2,108 (15.8%) died during hospitalization, yielding a case-fatality rate of 6.7% (95% CI: 6.4–7.0%). Higher mortality was independently associated with older age (adjusted Rate Ratio [aRR]: 2.3; 95% CI: 1.8–3.0), residence in the North region (aRR: 1.4; CI: 1.1–1.7)—a structurally disadvantaged area with poor health indicators—male sex (aRR: 1.3; CI: 1.2–1.4), lower household wealth (aRR: 1.3; CI: 1.1–1.6), Black (aRR: 1.2; CI: 1.0–1.4) and *Pardo* (Brown) (aRR: 1.2; CI: 1.0–1.3) race/ethnicity, and low schooling - illiterate people or people who have never attended school - (aRR: 1.2; CI: 1.0–1.5).

**Conclusions:** In-hospital AIDS mortality disproportionately affects individuals facing social vulnerabilities, including poverty, racial inequities, and geographic isolation. These findings highlight the enduring impact of structural inequities and reinforce the need for targeted social policies—such as cash transfer programs and expanded healthcare access—to reduce AIDS-related mortality in low- and middle-income countries.

**Funding:** This study was funded by the National Institute of Allergy and Infectious Diseases—NAIDS/NIH, Grant Number: 1R01AI152938.

## Introduction

Although expanded access to antiretroviral therapy (ART) has contributed to reducing HIV-related morbidity, mortality, transmission and, mortality, substantial social and regional disparities persist in Brazil.^1^ These inequalities create barriers to testing, early diagnosis, access and adherence to treatment among socially vulnerable populations living in the country’s poorest regions.^1^ Social disparities are recognized as key factors in the HIV/AIDS epidemic, which continues to affect, above all, economically disadvantaged populations with lower levels of schooling and belonging to the Black race/ethnicity.^2^ These determinants exert a profound influence on health outcomes, including an individual’s susceptibility to HIV infection, progression to AIDS, necessity for hospitalization, and risk of premature mortality. ^2^

Evidence from a literature review and meta-analysis suggested that hospitalizations due to complications associated with HIV infection—including opportunistic coinfections such as tuberculosis, cryptococcal meningitis, and severe bacterial infections— continue to be significant.^3^ People living with HIV (PLHIV) who require hospitalization are at a markedly higher risk of mortality. ^3–5^

Hospitalization data are essential for identifying priorities in case management and for improving the quality of life of patients with chronic conditions requiring lifelong treatment.^6^ However, severe morbidity among people living with HIV has been difficult to assess accurately, and data on its distribution and determinants over time remain limited.^6^ Observational hospital-based cohorts from Europe and North America have examined causes of hospitalization in the ART era as a proxy for severe morbidity, particularly in contexts of universal access to care.^6^ Updated information on the burden of severe disease can guide HIV clinicians in prioritizing screening and prevention strategies, as well as in monitoring patients in collaboration with specialists.^6^ Policymakers can also use such evidence to project future healthcare needs and associated costs.^6^ Previous studies have identified clinical predictors of hospitalization and in-hospital mortality,^6,7^ yet only one study^8^ has examined the social factors associated with these outcomes. Therefore, this study aimed to analyze the association between social determinants of health and in-hospital mortality among individuals living with HIV/AIDS.

## Methods

### Study design and data sources

This is a retrospective cohort study that covered 13,266 hospitalizations for AIDS from 2008 to 2018 in Brazil. The study population comes from the Cohort of 100 Million Brazilians, which corresponds to linked data from: i) *Cadastro Único* (CADU), a database that gathers records of people who apply for benefits from social programs; ii) records of hospitalizations for AIDS carried out in Brazil’s national health system known as the Unified Health System (SUS) and its Hospitalization Information System (SIH); and, iii) national mortality records extracted from the Mortality Information System (SIM). Because it is based on the CADU, the sample represents the poorest segment of the Brazilian population.

The data linkage process, previously validated and published,^9^ was carried out in two stages, the first being deterministic and the second probabilistic. The variables used to link the information are: name, date of birth, mother’s name and city of residence.^10^

People of both sexes and all ages who were hospitalized for AIDS between January, 1, 2008 and December, 31, 2018 were included in the study, excluding those with inconsistent dates, with less than 1 day of follow-up, and with missing information for the variables analyzed (Figure 1).

**Figure 1.**
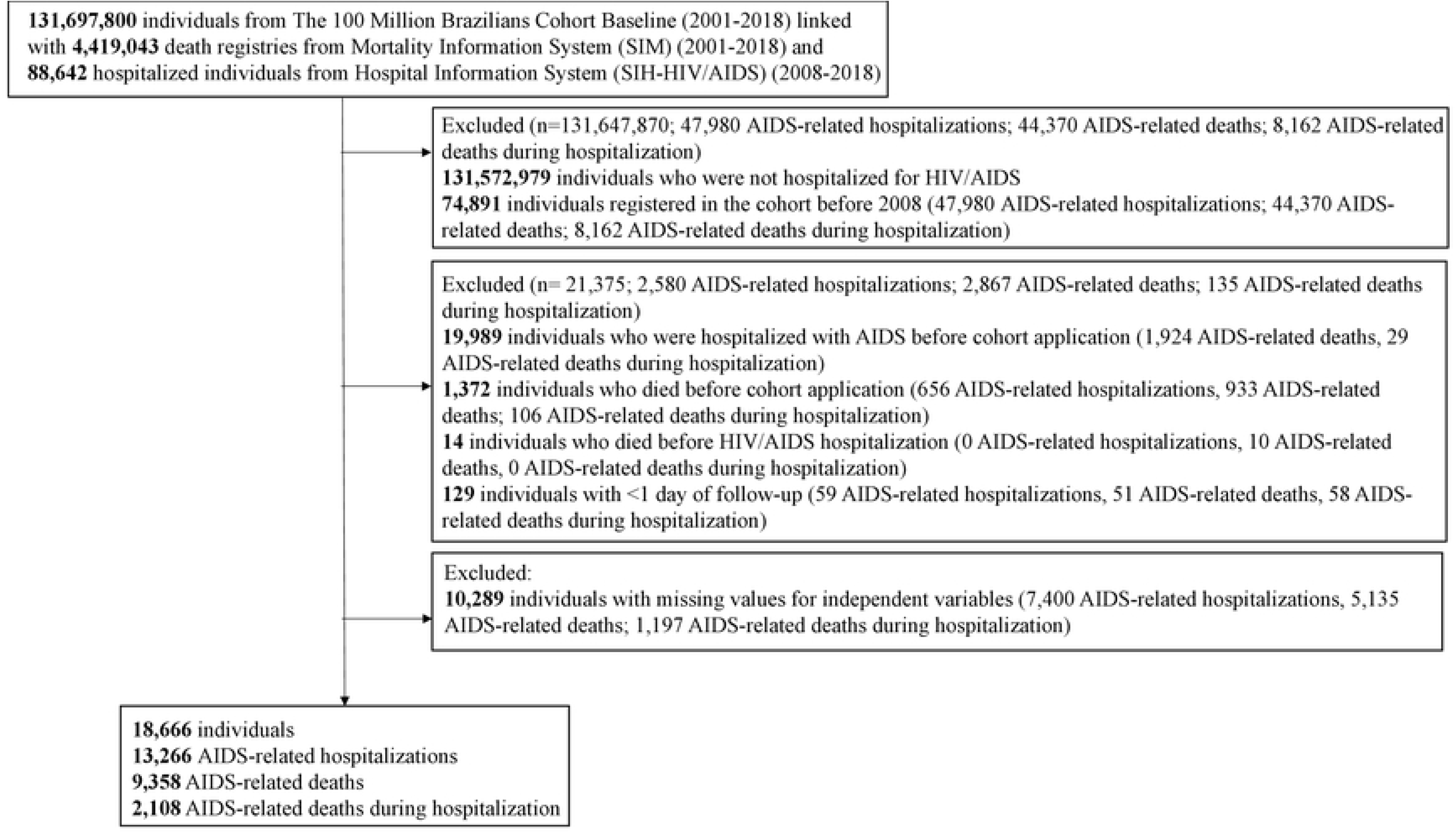
Flowchart selection of the study population, 2008-2018, Brazil.

The study protocol was approved by the Ethics Research Committee of the Institute of Collective Health of the Federal University of Bahia, under approval number 41691315.0.0000.5030 (Opinion n°: 3.783.920).

### Statistical analyses

Initially, a descriptive analysis was performed, using absolute numbers and percentages for categorical variables, according to the people who were hospitalized and according to the people who died from AIDS during hospitalization. In relation to numerical variables, an analysis of mean and standard deviation was performed for each subpopulation. The follow-up time in years (person-years at risk) was calculated starting on the date of entry into the cohort, until the first hospitalization for AIDS, death (whether due to AIDS or not) or end of follow-up (31 December 2018).

The primary outcome is death during hospitalization for AIDS, considering only each individual first AIDS-related hospitalization within the Brazilian hospitalization information system. AIDS-related death as the underlying cause, considering the international statistical classification of diseases and related health problems, 10th revision (ICD-10) codes related to AIDS (B20–B24). The outcome was calculated as a case fatality rate (%), defined as the number of people who died from AIDS during hospitalization divided by the total number of people in their first hospitalization for AIDS multiplied by 100 person-years at risk.

The analysis used a hierarchical approach^11^ without missing information, in which distal, intermediate and proximal independent determinants were assessed in blocks (Figure 2). Block 1 included the geographic variables: i) area of residence (rural or urban), and, ii) Brazilian region — North, Northeast, Central-West, Southeast and South — each defined by the Brazilian Institute of Geography and Statistics (IBGE). These regions group states with common physical, economic, cultural and social characteristics and serve as a basis for planning, statistical analysis and public policies, including in the area of health.^12^ The socioeconomic factors included in Block 2 were: i) self-declared race/ethnicity, which due to the small number of people of Asian origin were added to Whites, as they had similar income characteristics and social status. Regarding race/ethnicity, the “Pardo” category refers to individuals of mixed heritage, such as White and Indigenous, White and Black, Black and Indigenous, or Black and people of other colours or racial backgrounds. This group, like those classified as “Black,” is considered part of Brazil’s Afro-descendant population and, as a result, is likewise affected by the impacts of structural racism^13^; ii) education by years of schooling; and, iii) *per capita* household expenses in proportion to the minimum wage (MW), as a proxy for wealth and economic status. Block 3 included household infrastructure variables: i) house construction material; ii) water supply; iii) lighting; iv) sewage and, v) garbage disposal.

**Figure 2.**
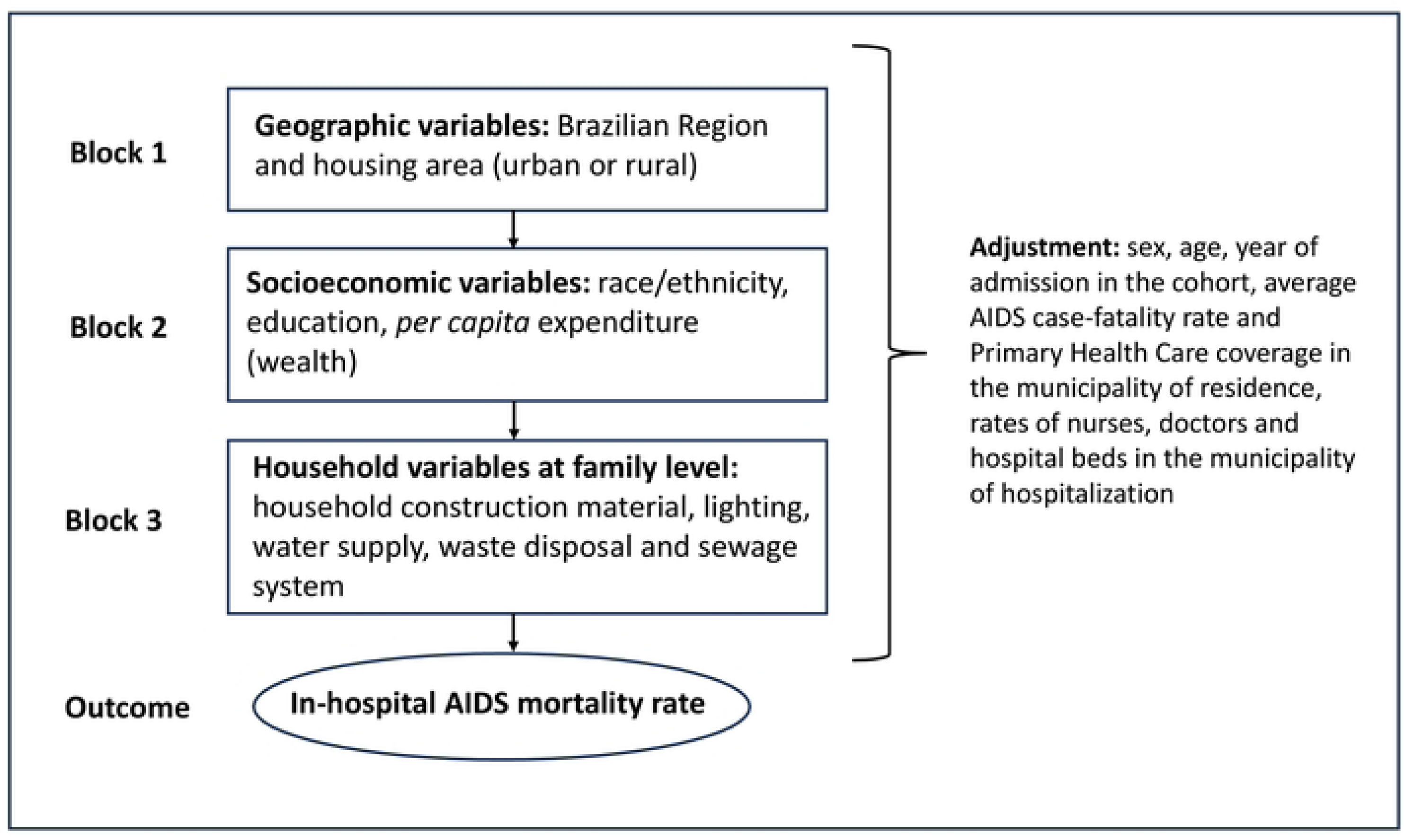
Hierarchical model of social determinants of health (SDOH) on in-hospital AIDS mortality rate.

The variables used for model adjustment were: age, sex, year of entry into the cohort, health variables related to the municipality of residence, such as Primary Health Care (PHC) coverage (%), and the average AIDS case-fatality rate, and hospital health variables related to the municipality of hospitalization, such as the rate of hospital beds, doctors and nurses per 1,000 inhabitants. All municipal variables were extracted from the Department of Informatics of the Unified Health System (DATASUS) database, which provides health information by municipalities in Brazil.^14^

We used multivariable Poisson regression models— as in similar SDOH studies—to estimate crude and adjusted associations for different SDOH with AIDS outcomes over the study period.^2,15^ Poisson regression models are widely used to analyze cohorts^16^ since they allow inclusion of ‘compensation/exposure’ terms (the time during which an individual is exposed to the risk of a certain outcome), and enable generation of incidence rate ratios (RR)^17^, which facilitate the interpretation of the results. Adjusted Poisson regression models were estimated using robust standard errors, clustered in the municipality of residence.

Model selection included retaining variables associated with in-hospital mortality from AIDS in each model, with a p-value of ≤0.10 for the partial models, and 0.05 for the final model, as in previous studies.^2^ Model 1 included factors specific to each analysis block (intra-block analysis). After filtering the variables in each specific block, backwards, stepwise regression was performed with hierarchical input from each block (inter-block analyses). Thus, Model 2 included variables from Blocks 1 and 2. Model 3 included final variables from Model 2 and Block 3. Some analyses of the final model were carried out according to sex groups and *per capita* household expenses according to the MW percentage.

To ensure the robustness of our findings, sensitivity analyses also tested alternative modeling approaches, including logistic, survival, negative binomial, and zero-inflated regression models (Supplementary Tables, pages 2-9).

The software STATA 15 Multiprocessor (MP) was used in all statistical analyses.

### Role of the funding source

The study funders played no role in the study design, data collection, data analysis, data interpretation, or writing of the paper. None of the authors were precluded from accessing study data, and they accepted the responsibility to submit for publication.

## Results

A total of 13,266 hospitalizations for AIDS were analyzed (Figure 1). Among these, 2,108 (15.9%) people died during hospitalization, yielding an in-hospital AIDS case-fatality rate of 6.7% (CI 95%: 6.4%-7.0%). The mean follow-up time per patient was 3,2 years. Regarding the main characteristics of hospitalized individuals, the mean length of stay was 15.2 days (SD 17.2), and the mean age was 37.3 years (standard deviation [SD]=12.5 years). Most participants were between 20 and 59 years old (10,918;82.3%), male (6,737; 50.8%), and of *Pardo (Brown)* race/ethnicity (6,923; 52.2%). A total of 5,231 (39.4%) reported having no income or an income equivalent to ≤ 0.24 of the minimum wage (MW), 9,744 (73.4%) had fewer than nine years of schooling, 12,578 (94.8%) lived in urban areas, and 5,192 (39.1%) resided in the Southeast region of Brazil. More than 85% of hospitalized people have adequate housing infrastructure, that is, they live in houses built with bricks, with water supply, lighting and garbage disposal provided by the public network, however, sewage collection carried out by the public network was lower, covering 8,334 people (62.8%) (table 1).

**Table 1.** Descriptive of independent variables by AIDS hospitalizations and AIDS-related deaths during hospitalizations, 2008-2018, Brazil.

| Independent variables | AIDS hospitalizations<br>N=13,266 | Deaths during<br>hospitalizations<br>N=2,108 |
| --- | --- | --- |
|  | n (%) | n (%) |
| Sex |  |  |
| Female | 6,529 (49.2) | 930 (44.1) |
| Male | 6,737 (50.8) | 1,178 (55.9) |
| Age |  |  |
| <19 years | 1,666 (12.6) | 232 (11.0) |
| 20-29 years | 3,019 (22.8) | 347 (16.5) |
| 30-39 years | 3,934 (29.6) | 631 (29.9) |
| 40-49 years | 2,611 (19.7) | 441 (20.9) |
| 50-59 years | 1,354 (10.2) | 283 (13.4) |
| >60 years | 682 (5.1) | 174 (8.2) |
| Race/ethnicity |  |  |
| White and Asian | 4,609 (34.7) | 698 (33.1) |
| *Pardo (Brown)* | 6,923 (52.2) | 1,131 (53.6) |
| Black | 1,697 (12.8) | 273 (12.9) |
| Indigenous | 37 (0.3) | 6 (0.3) |
| Years of schooling |  |  |
| >9 years | 3,522 (26.5) | 489 (23.2) |
| 5-9 years | 4,130 (31.1) | 595 (28.2) |
| Until 5 years | 4,652 (35.1) | 838 (39.7) |
| Illiterate/never went to school | 962 (7.2) | 186 (8.8) |
| Per capita household expenditures - % MW |  |  |
| >1 | 1,742 (13.1) | 232 (11.0) |
| 0.5-1 | 3,569 (26.9) | 565 (26.8) |
| 0.25-0.49 | 2,724 (20.5) | 419 (19.9) |
| 0-0.24 | 5,231 (39.4) | 892 (42.3) |
| Water supply |  |  |
| Public network | 11,339 (85.5) | 1,781 (84.5) |
| Well, spring or cistern | 1,927 (14.5) | 327 (15.5) |
| Home construction material |  |  |
| Brick | 11,274 (85.0) | 1,761 (83.5) |
| Wood or rammed earth | 1,992 (15.0) | 347 (16.5) |
| Lighting |  |  |
| Electricity | 12,725 (95.9) | 2,017 (95.7) |
| No electricity | 541 (4.1) | 91 (4.3) |
| Garbage disposal |  |  |
| Public network | 12,327 (92.9) | 1,961 (93.0) |
| Burned, buried or another | 939 (7.1) | 147 (7.0) |
| Sewage |  |  |
| Public network | 8,334 (62.8) | 1,319 (62.6) |
| Septic tank | 1,789 (13.5) | 304 (14.4) |
| Others | 3,143 (23.7) | 485 (23.0) |
| Brazilian Region |  |  |
| South-East | 5,192 (39.1) | 869 (41.2) |
| South | 2,649 (20.0) | 407 (19.3) |
| Mid-West | 1,049 (7.9) | 138 (6.5) |
| North-East | 3,147 (23.7) | 451 (21.4) |
| North | 1,229 (9.3) | 243 (11.5) |
| Area of residence |  |  |
| Rural | 12,578 (94.8) | 1,972 (93.6) |
| Urban | 688 (5.2) | 136 (6.4) |
| Year |  |  |
| 2008 | 1,759 (13.3) | 314 (14.9) |
| 2009 | 1,441 (10.9) | 239 (11.3) |
| 2010 | 1,721 (13.0) | 249 (11.8) |
| 2011 | 1,728 (13.0) | 290 (13.8) |
| 2012 | 2,468 (18.6) | 390 (18.5) |
| 2013 | 1,017 (7.7) | 153 (7.3) |
| 2014 | 1,212 (9.1) | 193 (9.2) |
| 2015 | 679 (5.1) | 109 (5.2) |
| 2016 | 551 (4.1) | 75 (3.6) |
| 2017 | 487 (3.7) | 72 (3.4) |
| 2018 | 203 (1.5) | 24 (1.1) |
| Municipal variables | Mean (SD) | Mean (SD) |
| Primary Health Care coverage* | 46.1 (28.6) | 45.8 (29.5) |
| Average AIDS case-fatality rate* | 27.7 (11.6) | 27.9 (12.2) |
| Hospital beds for 1,000 inhabitants** | 2.6 (1.5) | 2.5 (1.5) |
| Nurses for 1,000 inhabitants** | 0.9 (0.4) | 0.8 (0.4) |
| Doctors for 1,000 inhabitants** | 2.0 (1.2) | 1.8 (1.2) |

MW=minimum wage. SD=standard deviation
\*In the municipality of residence \*\*In the municipality of hospitalization

Among those who died during hospitalization the mean length of stay for the hospitalizations was 16.8 days (SD 18.0). In this population, we observed a higher percentage of men (55.9%, n=1,178), with a higher mean age (40.4 years, SD=12.6) and a higher percentage of people over 60 years of age (8.2%; n=174). There was also a higher percentage of *Pardo (Brown)* people (53.6%, n=1,131), with up to 9 years of schooling (n=1,619, 76.8%), with low *per capita* expenses (42.3%, n=892). In the population that died during hospitalization, we noted a slightly higher proportion of people living in the Southeast (n=869; 41.2%) and North (n=243; 11.5%) regions of Brazil. Housing characteristics are very similar to those of hospitalized people (table 1).

Regarding geographic factors, people living in the North region of Brazil had a higher risk of dying from AIDS during hospitalization (adjusted Rate Ratio [aRR]: 1.37; 95% CI: 1.13-1.67). The socioeconomic factors associated with in-hospital AIDS mortality were: *per capita* household expenditures corresponding to 0 to 0.24 of the MW (aRR: 1.30; 95% CI: 1.06-1.59), Black (aRR: 1.21; 95% CI: 1.02-1.43) or *Pardo (Brown)* (aRR: 1.17; 95% CI: 1.04-1.32) race/ethnicity, and, up to 5 years of schooling (aRR: 1.17; 95% CI: 1.01-1.36). In the final model, there were no household characteristics associated with in-hospital AIDS mortality. Regarding the adjustment variables, men showed an association with in-hospital mortality (aRR: 1.29; 95% CI: 1.17-1.41), as well as the oldest people in the sample (50 to 59 years-old [aRR: 1.67; 95% CI: 1.30-2.13] and over 60 years old [aRR: 2.32; 95% CI: 1.80-2.99]) (Table 2).

**Table 2.**
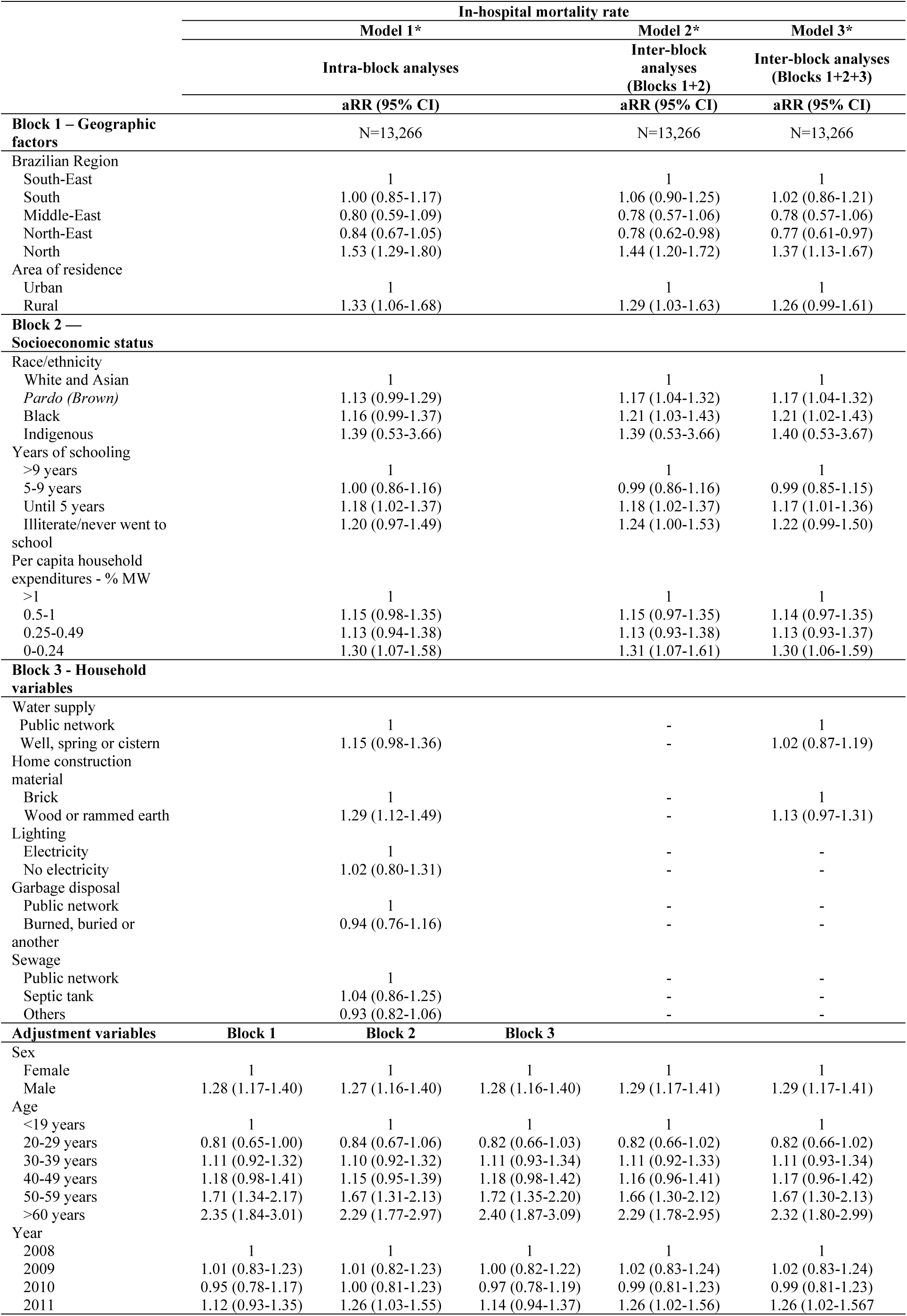
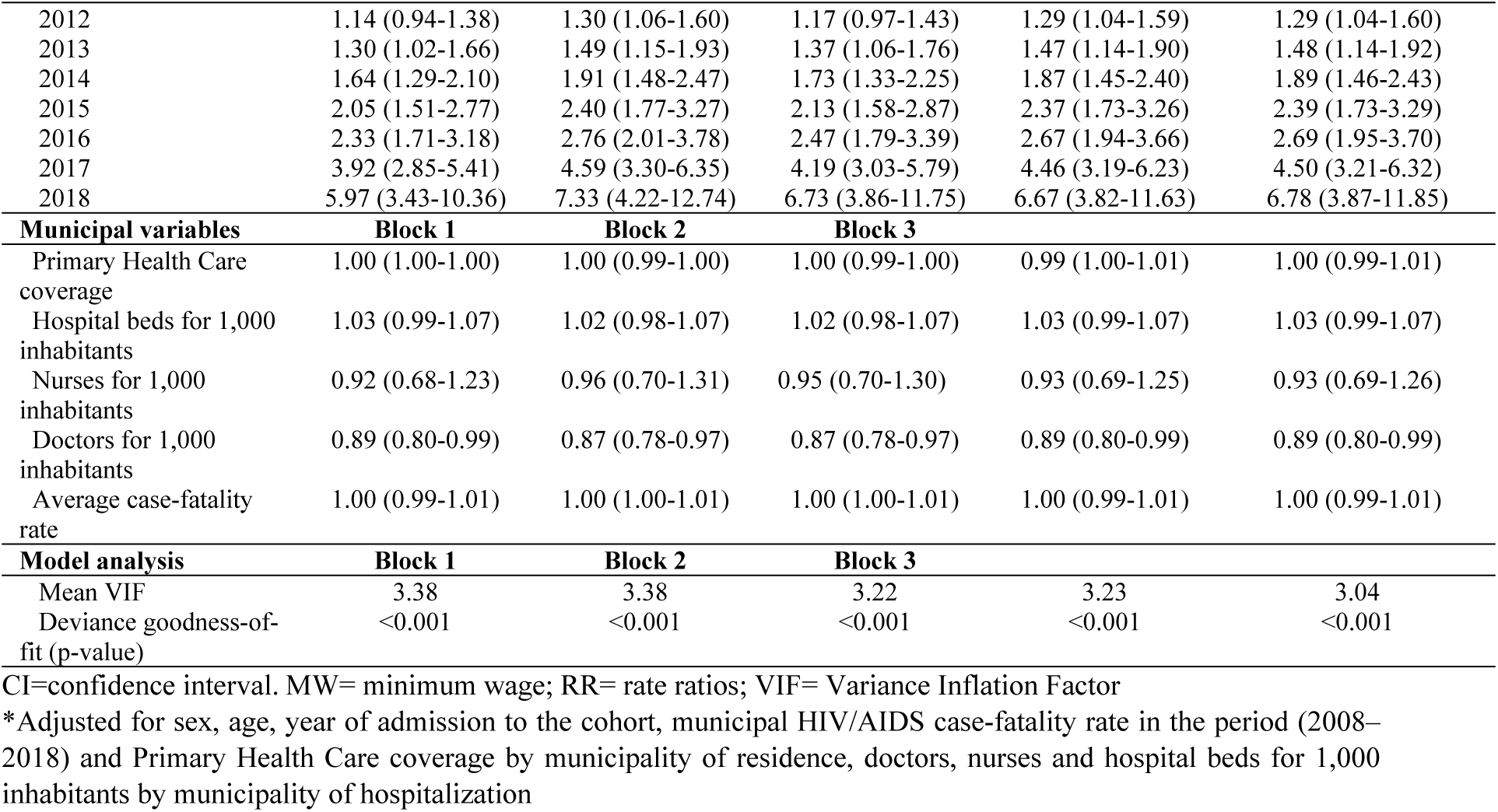
Effect of social determinants of health on AIDS in-hospital mortality rate in Brazil, results of Hierarchy Analysis using multivariable Poisson Regression, 2008-2018

Among women, there were no geographic or household factors associated with AIDS mortality during hospitalization. However, there were associated socioeconomic factors, such as: being *Pardo (Brown)* (aRR: 1.36; 95% CI: 1.14-1.61) or Black (aRR: 1.24; 95% CI: 1.00-1.55), or having any lower category of *per capita* expenditure. Among men, the North Region demonstrated an association with in-hospital mortality (aRR: 1.43; 95% CI: 1.13-1.80); as well as Black (aRR: 1.30; 95% CI: 1.09-1.54) or Indigenous (aRR: 1.35; 95% CI: 1.02-1.79) race/ethnicity and low schooling level (up to 5 years of study [aRR: 1.30; 95% CI: 1.09-1.54) and illiterate or never went to school [aRR 1.35; 95% CI 1.02-1.79]). No household characteristic was associated with risk of in-hospital death among males (Table 3).

**Table 3.**
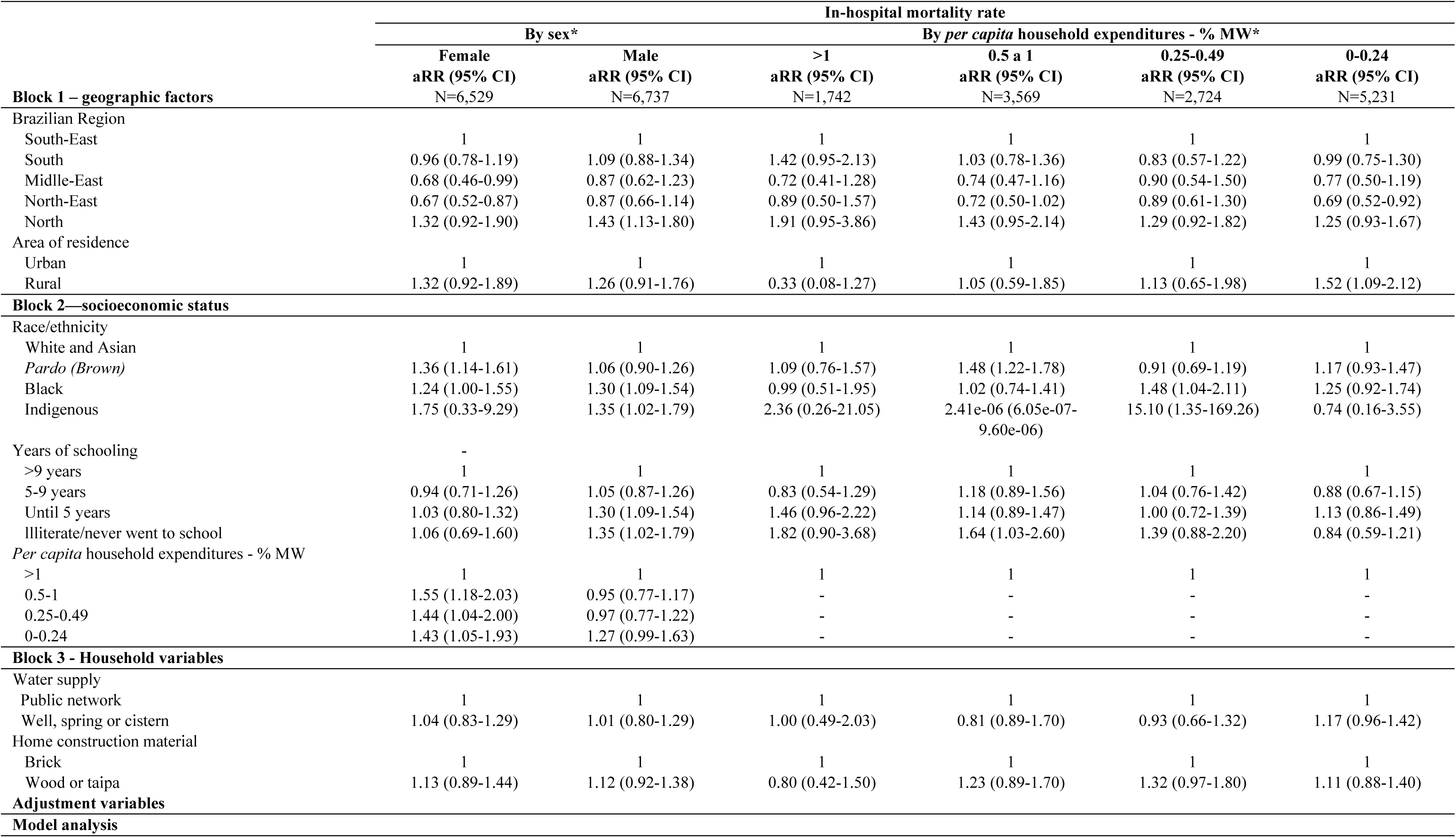
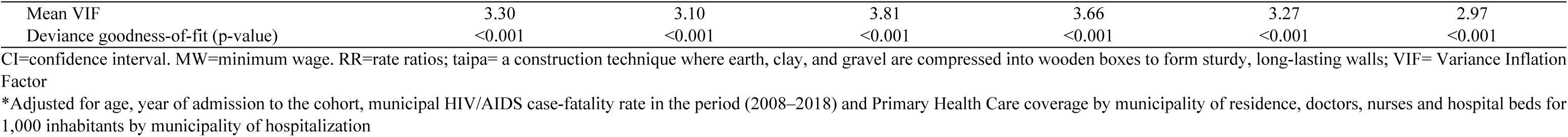
Effect of social determinants of health on AIDS in-hospital mortality rate in Brazil, results of Hierarchy Analysis using multivariable Poisson Regression by sex and by *per capita* household expenditures, 2008-2018

Among those living in extreme poverty, that is, those who have no or up to 0.24% of *per capita* expenses in relation to the minimum wage, living in rural areas was associated with in-hospital mortality from AIDS (aRR: 1.52; 95% CI: 1.09-2.12). On the other hand, among those individuals with higher income, none of the factors analyzed was associated with in-hospital mortality due to AIDS (table 3).

## Discussion

To our knowledge, this is the largest study that evaluated the association between social determinants of health (SDOH) and in-hospital mortality from AIDS or other infectious diseases. In this individual-level cohort study, we identified 13,266 hospitalizations due to AIDS over an 11-year period and analyzed the SDOH associated with in-hospital death. Key geographic factors—such as residence in the poorest regions of the country—and social factors, including identification as *Pardo (Brown)* or Black, low educational attainment, and poverty, were significantly associated with in-hospital mortality. Demographic characteristics, such as being male and aged over 50 years, were also linked to higher risk of death during hospitalization.

The in-hospital mortality rate we found in this study (6.7%) was similar to that found in a previous cohort study carried out in a hospital in Rio de Janeiro, Brazil, (6.6%),^6^ although it was less than half the rate of a study conducted in Malawi (14.6%), a sub-Saharan African country with one of the highest prevalence of HIV/AIDS in the world and limited hospital resources.^18^

The SDOH most strongly associated with in-hospital mortality was a geographic factor, that is, living in the North of the country, a region that, for geopolitical reasons, has lower economic and social development compared to other regions of the country,^19^ which may imply less access to healthcare, late diagnosis and adherence to treatment. The causes of difficulty in adhering to ART may be due to individual factors, social and economic factors, access to the health system, lack of integrated care, and structural and political factors such as living far from treatment centers.^20^ In this sense, living in marginalized regions, such as the northern region of Brazil, is a factor that interferes with adherence. These social vulnerabilities are not only markers of systemic exclusion, but also key drivers of adverse health outcomes such as higher mortality from AIDS.^2^

According to national data, between 2012 and 2022, the North Region showed an increase in the standardized mortality rate due to AIDS.^20^ This region has the lowest rates in the country related to the cascade of continuous HIV care for the year 2021.^1^ Namely, the five federative units of the northern region presented a proportion of 83 to 89% of people living with HIV/AIDS linked to some health service; among these people, the proportion of ART completion ranged from 79 to 85% and 70 to 74% presented viral suppression (viral load <50 copies/mL).^1^ These same rates demonstrate better results in the federative units of the Southeast region, which has the greater economic development in the country, that is, 91 to 92%; 87 to 89% and 78 to 81% respectively.^1^ These disparities illustrate how regional inequality directly impacts health outcomes and the effectiveness of public health strategies as well as the perception of risk for HIV^21^. Such differences are emblematic of the broader structural inequalities in a country as vast and diverse as Brazil, with its continental dimensions.^22^

In-hospital mortality was higher in the Black and *Pardo (Brown)* population, which also have a strong association with the incidence and mortality from AIDS.^2^ Nationwide data demonstrated that, in 2022, 61.7% of deaths occurred among Black and *Pardo (Brown)* people (47.0% in *Pardo* and 14.7% in Black people).^20^ Comparing the years 2012 and 2022, there was a decrease of 1.6% in the number of deaths among Black people, however, deaths increased by 12.0% among *Pardo (Brown)* people.^20^ Clinical data such as people linked to some health service, undergoing ART and viral suppression, timely initiation of ART (with CD4 ≥500 cells/mm³) and adherence indicators were always more unsatisfactory among Black and *Pardo (Brown)* people than among White and Asian people.^13^ These negative results occur because, in Brazil, Black and *Pardo (Brown)* people are subject to structural and institutional racism, which is deeply rooted in systems, laws, and policies that perpetuate widespread unfair treatment and oppression, imposing worse access to education, decent work, adequate income and healthcare, generating harmful consequences for health.^13^

Individuals with low levels of schooling were more likely to die during hospitalization for AIDS, and also had a higher incidence, mortality, and case-fatality rate due to the disease.^2^ In general, people with few years of formal education face more difficulties in receiving and understanding health information, as well as in transforming it into practical actions for their well-being. In addition, they are more vulnerable to risk behaviors, late diagnosis of HIV/AIDS, no access to health services, and untimely initiation and adherence to ART.^1,2^

Low economic status has also been associated with in-hospital death from AIDS, as well as maintaining an association with high incidence and mortality from the disease.^2^ People in an adverse economic conditions are prone to inadequate nutrition, in addition to living with social marginalization that acts as a barrier to access to health services, as well as to early diagnosis and satisfactory linkage to health services and early treatment of HIV/AIDS.^2^

As expected, people over 50 years of age had higher in-hospital mortality from AIDS, a result also found in a previous study conducted in Rio de Janeiro.^6^ Overall mortality from AIDS in Brazil is also higher in this age group.^20^ This is probably because people with HIV/AIDS are living longer which makes them more vulnerable to serious co-morbid events.^6^

Although in Brazil men have better adherence to ART than women, we found,^1^ we found higher in-hospital mortality due to AIDS among men, similar to a study carried out in Malawi.^18^ An observational study conducted in Myanmar found that the only social determinant associated with AIDS-related hospitalization and death was male gender.^8^ These results reinforce the role of the male gender in negative outcomes due to HIV/AIDS. Men may take longer to seek hospital care, or may be at greater risk for advanced infection and malignancies, the consequence of which is worse prognosis.^20^

One study conducted in the United States demonstrated that although hospitalization improved HIV clinical outcomes such as viral suppression, appointment attendance, retention in care, and decreased acute care utilization, a nurse-led hospital-to-community transition program was able to address social needs and contribute more positively to these outcomes.^23^ People living with HIV/AIDS in Brazil could benefit from programs like this, which strengthen the hospital’s link with primary care services or other specialized networks.

Considering the importance of the social determination of infectious diseases in Brazil, in 2024, the *Programa Brasil Saudável* (Healthy Brazil Program) was launched, program coordinated by the Ministry of Health that joins efforts of several other ministries and partners of the federal government to expand actions with the objective of eliminating these infectious diseases as public health problems in Brazil by 2030, including AIDS and vertical transmission of HIV, in alignment with the Sustainable Development Goals of the United Nations.^24^ One of the strategies of the *Programa Brasil Saudável* is to enhance the financial and technical sustainability for managing the continuum of care for HIV and AIDS, taking into account regional differences in the behavior of the epidemic and the effectiveness of the programmatic response.^24^ Two main objectives are: to expand and qualify the provision of diagnostics and linkage strategies related to HIV and AIDS, throughout the national territory, prioritizing the most vulnerable populations; and to expand access to comprehensive care, aiming at improving retention and adherence to treatment for people living with HIV and AIDS.^24^

In fact, expanding access to ART is a central component of providing quality care to people living with HIV/AIDS and preventing spread of the epidemic.^18^ Despite significant progress in universalizing ART, challenges persist, especially for vulnerable populations.^25^ Economic vulnerability is a critical determinant of adherence to HIV treatment.^25^ Individuals facing poverty, unemployment, or food insecurity often encounter barriers to consistently accessing and adhering to ART regimens, including transportation costs, competing daily survival priorities, and limited health literacy.^25,26^ These challenges highlight the need for integrated responses that go beyond biomedical interventions.^26^ In this context, the expansion of social protection policies—especially income transfer programs—has proven to be a valuable complement to ART access strategies, since they have demonstrated at the individual and ecological levels their potential to reduce incidence, hospitalization and mortality from AIDS, among the poorest individuals.^25–27^

It is important to underscore that stigma functions as a critical social determinant of health and is closely linked to adverse treatment outcomes among people living with HIV/AIDS^28^. Although stigma was not directly measured in the present study—partly due to the inherent complexity of doing so—the strong association between stigma and social vulnerability allows for reasonable inference.^28^ The elevated in-hospital mortality observed in the North region, as well as among Black and mixed-race individuals with lower levels of education and income, is very likely influenced by stigma embedded in the care provided to these populations.

Despite being an unprecedented study that provides a novel perspective on AIDS-related hospitalizations, our analysis has some limitations. It is restricted to low-income individuals registered in the CADU and admitted to public hospitals within Brazil’s Unified Health System (SUS). To ensure data reliability, individuals with missing information for any study variable were excluded. Another limitation is the absence of detailed clinical and hospital-level variables. However, to minimize potential bias, we incorporated contextual indicators such as healthcare workforce density (number of physicians and nurses per 1,000 inhabitants) and hospital infrastructure (number of hospital beds per 1,000 inhabitants) in the municipality where each hospitalization occurred.

In conclusion, despite universal access to antiretroviral therapy (ART) and a public healthcare system free of charge at the point-of-care, key social determinants of health (SDOH)—such as geographic, socioeconomic, and racial factors—remain associated with higher in-hospital mortality from AIDS in Brazil. Structural inequalities in healthcare access continue to drive adverse outcomes, particularly among individuals living in poorer regions, those identifying as Black or *Pardo*, and people with low schooling or extreme poverty.^25,26^ Policies such as conditional cash transfers, educational initiatives, and regionally tailored healthcare strategies can reduce disparities, improve treatment adherence, and enhance survival. Strengthening health equity in low- and middle-income countries (LMICs) therefore requires integrated policies that address social and structural roots of inequality, targeting vulnerable groups.

## Author contributions

PFPSG= conceptualization, formal analysis - accessed and verified the data, investigation, methodology, decision to submit the manuscript, writing – original draft, and writing– review & editing

GSJ= formal analysis - accessed and verified the data, methodology and writing– review & editing

AFS= methodology, writing– review & editing IL= methodology, writing– review & editing CAST= writing– review & editing

LES= project administration, funding acquisition, writing– review & editing JM= project administration, funding acquisition, writing– review & editing ID= project administration, funding acquisition, writing– review & editing

DR= conceptualization, project administration, funding acquisition, investigation, methodology, supervision - accessed and verified the data, decision to submit the manuscript, writing– review & editing

## Data sharing statement

The data underlying this article will be made available on reasonable request to the Center for Data and Knowledge Integration for Health (CIDACS), Oswaldo Cruz Foundation (Fiocruz), subject to ethical approval and compliance with applicable data protection and governance requirements.

## Declaration of interests

We declare no competing interests.

I, Priscila FPS Gestal, as corresponding author, confirm that all authors have seen and approved of the final text.

## Acknowledgement section

Funding: This study was funded by the National Institute of Allergy and Infectious Diseases—NAIDS/NIH, Grant Number: 1R01AI152938. The funders had no role in the study design or in the decision to submit this manuscript.

ChatGPT (OpenAI) and Gemini (Google) was used solely to assist with improving the English language and clarity of selected sections of the Introduction and Discussion. The authors take full responsibility for the content of the manuscript.

## Notes

### Competing Interest Statement

The authors have declared no competing interest.

### Funding Statement

Yes

### Author Declarations

The study protocol was approved by the Ethics Research Committee of the Institute of Collective Health of the Federal University of Bahia, under approval number 41691315.0.0000.5030 (Opinion nr: 3.783.920).

